# A Robust Cell-Free RNA Approach for the Early Detection of Colorectal Cancer

**DOI:** 10.64898/2026.07.01.26357015

**Authors:** Pablo Monteagudo-Mesas, Laura Sánchez, Giovanni Asole, Beatriz Neto, Cristina Tuñí-Domínguez, Lucila González, Elena Cristina Rusu, Lluc Cabús, Sonia Panadero-Fajardo, Purificación Catalina, Sol Garcia, Pilar Simón-Extremera, Laura Padilla García, Julien Lagarde, Phil Sanders, Marc Weber

**Affiliations:** Flomics Biotech, SL, Carrer de Roc Boronat 31, 08005 Barcelona, Spain; Centre for Genomic Regulation (CRG), The Barcelona Institute of Science and Technology, Dr. Aiguader 88, Barcelona 08003, Spain; Universitat Pompeu Fabra (UPF), Barcelona, Spain; Andalusian Public Health System Biobank, Coordinating Node, Av. del Conocimiento, S/N, 18016 Granada, Spain; Health and Biomedicine, Leitat Technological Center, Barcelona Science Park, Barcelona 08028, Spain

## Abstract

Colorectal cancer (CRC) screening remains limited by patient adherence and sub-optimal sensitivity for early-stage disease. While liquid biopsy has revolutionized cancer diagnostics, cfDNA-based methods often struggle with early-stage detection due to low analyte levels. Here, we present a robust cell-free RNA (cfRNA) platform for the early detection of CRC. Using a retrospective cohort of 255 healthy controls and 250 CRC patients, we implemented an optimized workflow featuring a RUVg-based normalization strategy to remove platelet-driven transcriptomic noise. We identified differentially expressed genes enriched in key CRC-associated biological pathways, including inflammation, EMT, and metabolic dysregulation. An XGBoost classifier trained on these features achieved a mean AUC of 0.92 in cross-validation and 0.89 in a validation cohort, demonstrating 67% sensitivity at 90% specificity. Notably, our platform showed particular efficacy in identifying early stage cancer (stage I and II), achieving 73.7% sensitivity at 90% specificity. These findings suggest that cfRNA profiling offers a powerful, non-invasive orthogonal approach to CRC screening, capable of overcoming the sensitivity limitations of DNA-based assays in early-stage disease.

## Introduction

Colorectal cancer (CRC) is the third most commonly diagnosed cancer and the second leading cause of cancer-related death worldwide, accounting for over 1.9 million new cases and 900,000 deaths annually (Bray *et al*., 2024). Incidence and mortality are projected to increase by 70% over the next 20 years, and this trend is compounded by an increase in early-onset CRC among adults under 50 (Saraiva, Rosa and Claro, 2023). Crucially, survival in CRC is highly stage-dependent: 5-year survival exceeds 90% for localised stage I disease (Cardoso *et al*., 2022), but falls to 14% once distant metastases are present (Shin, Giancotti and Rustgi, 2023). This steep stage effect, together with the considerable toxicity of the systemic therapies used against advanced disease (Letai and de The, 2025), makes the detection of CRC before dissemination one of the most promising approaches for improving outcomes and survival.

Organised screening programmes have been instrumental in reducing CRC incidence and mortality through the detection and removal of precursor lesions. Colonoscopy remains the reference modality, while faecal immunochemical testing (FIT) provides a less invasive, population-scalable alternative (Quintero *et al*., 2012). Despite the success of these programmes they are subject to substantial limitations such as patient non-adherence (Ascunce *et al*., 2010) and sub-optimal sensitivity for early-stage lesions (Kim *et al*., 2017; Niedermaier, Balavarca and Brenner, 2020). Due to these issues and the concerning increase in CRC incidence among younger adults, the development of highly sensitive, specific, and non-invasive detection methods is essential.

Liquid biopsy has emerged as a highly promising approach for cancer early detection, including CRC, as it allows minimally invasive and repeatable analysis of circulating tumour-derived analytes from biofluids including blood and urine (Batool *et al*., 2023). Although many analytes are available for analysis by liquid biopsy, including cell-free RNA and DNA, circulating tumour DNA, circulating tumour cells, and proteins and other analytes, to date clinical translation in CRC has been dominated by cell-free DNA-based methods including somatic mutation detection (Strickler *et al*., 2018), fragmentomics (Bruhm et al., 2025), and methylation (Lin et al., 2026, Liu et al., 2020, Moore et al., 2025).

While great successes have been achieved with cfDNA-based methods (Onieva-García et al., 2015, Chung et al., 2024), these approaches are ill-suited to detect early stage cancer due to the limited amount of DNA released into the bloodstream and other biofluids during cell death (Haque and Elemento, 2017). As a result cfDNA-based approaches display reduced sensitivity for early-stage cancer detection (Liu et al., 2020; Gao et al., 2022).

In recent years cell-free RNA (cfRNA) has emerged as a promising alternative to cfDNA in liquid biopsies. cfRNA is present in all biofluids and is composed of a diverse array of species including protein-coding mRNAs, long non-coding RNAs, small RNAs, and circular RNAs (X. Wang *et al*., 2025). While cfRNA is released during cell death as occurs with cfDNA, it is also actively secreted by live cells. Importantly, the content of the RNA released into circulation depends on the state of the cell (Vorperian, Moufarrej and Quake, 2022a; Stejskal *et al*., 2023). Whereas cfDNA primarily reflects the relatively static genomic alterations of tumour cells, cfRNA provides a dynamic readout of tumour-associated dysregulation, capturing aberrant processes such as altered gene expression and alternative splicing. cfRNA also captures transcriptional changes both in the tumour microenvironment and at the systemic level as the body responds to the tumour (Kan *et al*., 2023; Kim and Yoo, 2026). As a result, early changes that occur in cancer can potentially be detected with cfRNA before detection can be achieved with cfDNA.

Due to these properties, cfRNA is uniquely positioned to capture subtle, early changes in cancer that are not visible to DNA-based assays (Kim and Yoo, 2026). This potential is being realised with a growing number of studies demonstrating the value of the cell-free transcriptome for cancer detection. Early work provided the first transcriptome-wide characterisation of cfRNA in cancer, identifying tissue- and cancer-specific genes that were recurrently detected in patients with cancer but largely absent in non-cancer individuals (Larson *et al*., 2021). Building on this, cfRNA-based classifiers have since achieved strong discriminatory performance across multiple cancer types: the combination of human and microbe-derived plasma cfRNAs distinguished cancer patients from healthy donors across five cancer types (Chen *et al*., 2022), while more recent blood-based circulating RNA classifiers have reported promising classification accuracy for several tumour types, including colorectal cancer (F. Wang *et al*., 2025; Momen-Roknabadi *et al*., 2025). Together, these advances demonstrate that cfRNA carries disease-specific information that is detectable in a minimally invasive manner, making it a highly promising analyte for cancer early detection including CRC.

Despite its great potential, the clinical utility of cfRNA is challenged by significant technical and biological obstacles. The inherently low concentration and rapid degradation of cfRNA in plasma require rigorous experimental and bioinformatic pipelines to generate high-quality data (Cabús *et al*., 2022). Key challenges include the management of pre-analytical variables, which can introduce systematic biases (The exRNAQC Consortium *et al*., 2025; Tuñí-Domínguez *et al*., 2026), and the pervasive presence of “unwanted variation”—most notably from platelet-derived transcripts—which can confound the detection of disease-specific signals (Nesselbush *et al*., 2025). In the absence of standardised sample processing protocols and quality-control criteria, these factors compromise reproducibility and hinder comparison across studies. Achieving robust, reproducible cfRNA profiles in such a noisy, low-input regime, and at the scale required for biomarker discovery, is therefore a prerequisite for moving cfRNA assays toward clinical application.

In this study, we address these challenges by developing and implementing an optimized cfRNA experimental and analytical workflow, which we apply to a large retrospective cohort of colorectal cancer patients and healthy controls. By combining a standardised sample processing protocol with stringent quality-control filtering and a robust platelet-correction strategy, we establish a platform that delivers high-quality cell-free transcriptome profiles reproducibly and at scale. We show that this approach distinguishes colorectal cancer patients from healthy controls, and identifies differentially expressed genes that are enriched in biological pathways with well-established roles in CRC, confirming that the captured signal reflects genuine tumour-associated biology rather than technical noise. Building on these features, a machine learning classifier accurately separates CRC from healthy control samples, achieving a performance for early stage cancer (stage I and II) of 73.7% sensitivity at 90% specificity, highlighting its potential to detect disease at a clinically actionable stage. Together, these results establish cfRNA as a promising, non-invasive analyte for colorectal cancer detection and provide a foundation for the application of cfRNA in multi-cancer early detection.

## Results

### Study design and workflow

We obtained a retrospective cohort of healthy control individuals (n=255) and colorectal cancer (CRC) patients (n=250) from Spanish biobanks to evaluate the potential of plasma cell-free RNA (cfRNA) as a non-invasive diagnostic biomarker (Figure 1A). To ensure robust model development and to provide an unbiased estimate of generalization on unseen data, the total patient population was partitioned into a training cohort (70%) for feature selection and model development, and an internal validation cohort (30%) for final performance evaluation (Figure 1F). Demographic analysis confirmed that age (Figure 1C) and cancer stage (Figure 1B) distributions were closely matched across both cohorts. We observed a higher proportion of males in the CRC group compared to the healthy controls (Figure 1D); this demographic imbalance was accounted for by including patient sex as a covariate in all downstream differential expression analyses to prevent confounding.

**Figure 1.**
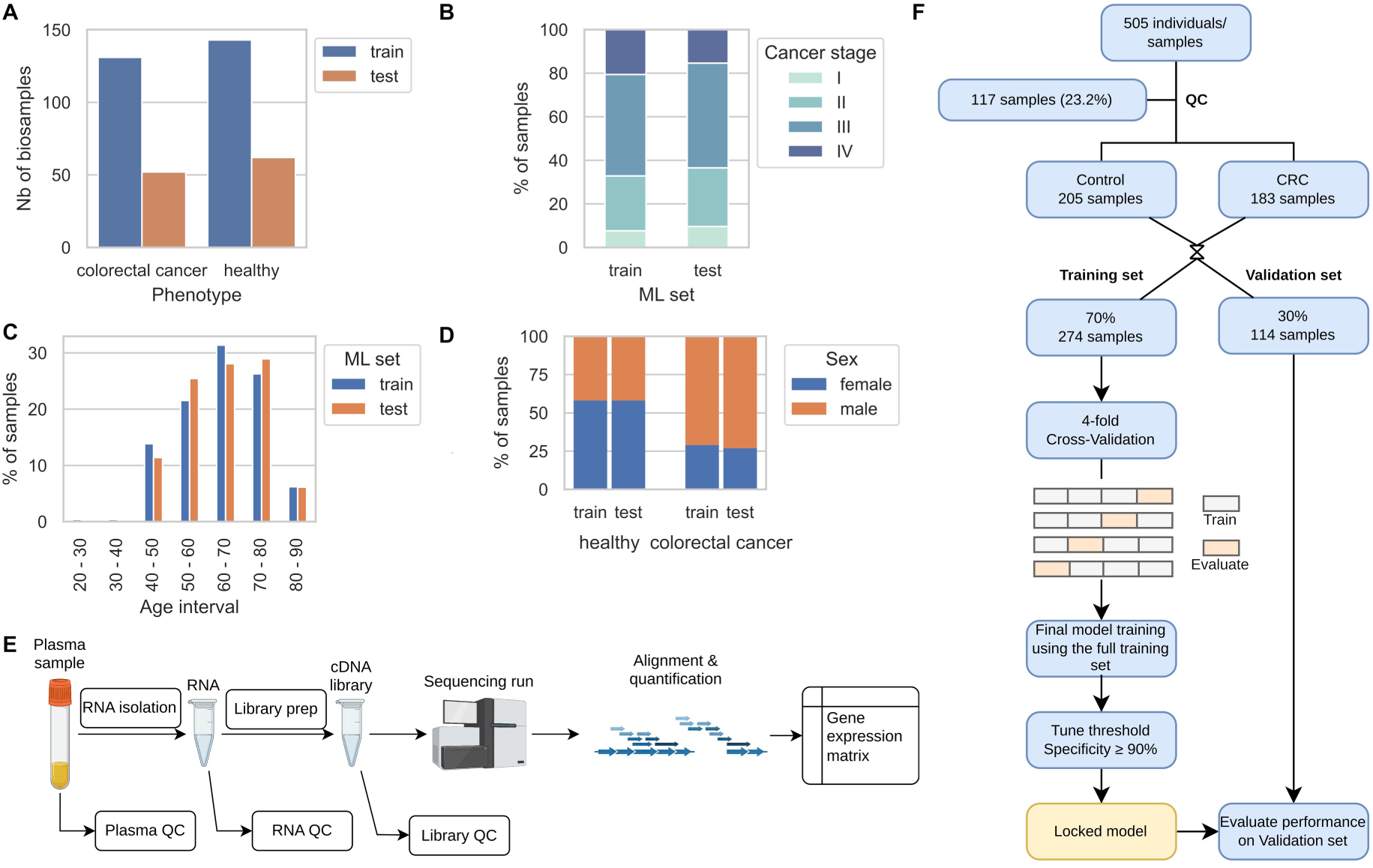
Overview of the cohorts composition, cfRNA analysis pipeline and machine learning (ML) workflow. Overview of the training and validation cohorts composition, after QC filtering. (**A**) Number of colorectal cancer (CRC) and healthy control samples, in the training and the validation cohorts. (**B-D**) Number of samples in the training and in the validation cohorts, broken down by (**B**) cancer stage, (**C**) age interval, and (**D**) sex. (E) Overview of the optimized cfRNA experimental protocol and cfRNA-seq data preprocessing workflow, from plasma sample to gene expression matrix. (F) Overview of the study design and the ML workflow. A total of 505 individuals were included based on the inclusion and exclusion criteria. Plasma samples for those were processed using our optimized cfRNA pipeline. Quality control based on the cfRNA-seq data quality metrics excluded 117 samples (23.2%). Of the remaining 388 samples, 114 were reserved for the validation cohort, whereas the training cohort was used to perform cross-validation and training, and lock the ML model. The locked model was finally evaluated on the validation cohort.

The overall workflow of this study encompassed three main phases: an optimized cfRNA experimental sample processing protocol (Figure 1E), a bioinformatics data preprocessing pipeline with a cfRNA-specific QC module (including gene quantification, blacklisting, and normalization), and a machine learning workflow (Figure 1F). The machine learning pipeline utilized cross-validation strictly on the training set for feature selection and hyperparameter tuning before final evaluation on the validation set.

### Optimized cfRNA platform delivers high quality cell free transcriptome profiles

To address the inherently low concentration and fragmented nature of plasma cfRNA, we implemented an optimized experimental protocol featuring a double DNase treatment. This step rigorously minimized genomic DNA (gDNA) contamination, a well-documented technical artifact that can severely skew cfRNA quantification (Tuñí-Domínguez *et al*., 2026). We processed and sequenced 505 plasma samples across the training and validation cohorts using our optimized experimental protocol. Following sequencing, the libraries exhibited a median depth of 32.89 million reads. Our optimized protocol successfully captured a diverse and complex cell-free transcriptome, detecting a total of 10,017 genes above a threshold of 10 reads in at least 80% of the samples.

The analysis of cell-free transcriptomes is highly sensitive to technical noise and pre-analytical variables, such as sample handling, plasma isolation protocol, and storage (Kim et al., 2022; The exRNAQC Consortium et al., 2025). Therefore, rigorous quality control (QC) is essential to exclude low-quality libraries that could compromise downstream biomarker discovery. Given that experimental standardization alone cannot fully mitigate variability arising from pre-analytical conditions, we further assessed sample quality using the sequencing data to identify and exclude low-quality libraries. To define the set of high-quality cfRNA profiles we applied three stringent QC filters. First, we required a minimum sequencing depth of 2 million exonic reads to remove failed libraries, samples with insufficient input material, or highly contaminated samples (Figure 2A, Figure S1). Second, to further avoid genomic DNA (gDNA) contamination, we required the fraction of spliced reads (FSR) to exceed 20%. Third, because library diversity dictates the probability of detecting rare circulating biomarkers, we enforced a threshold requiring an NG80 metric (the number of genes accounting for 80% of all mapped reads) of at least 500. This metric ensured that our expression profiles were broad and not artificially dominated by a few highly abundant transcripts (Tuñí-Domínguez et al., 2026). By applying these stringent QC thresholds, we identified a set of 388 high-quality samples that were used in all the downstream analyses (Figure 1F).

**Figure 2.**
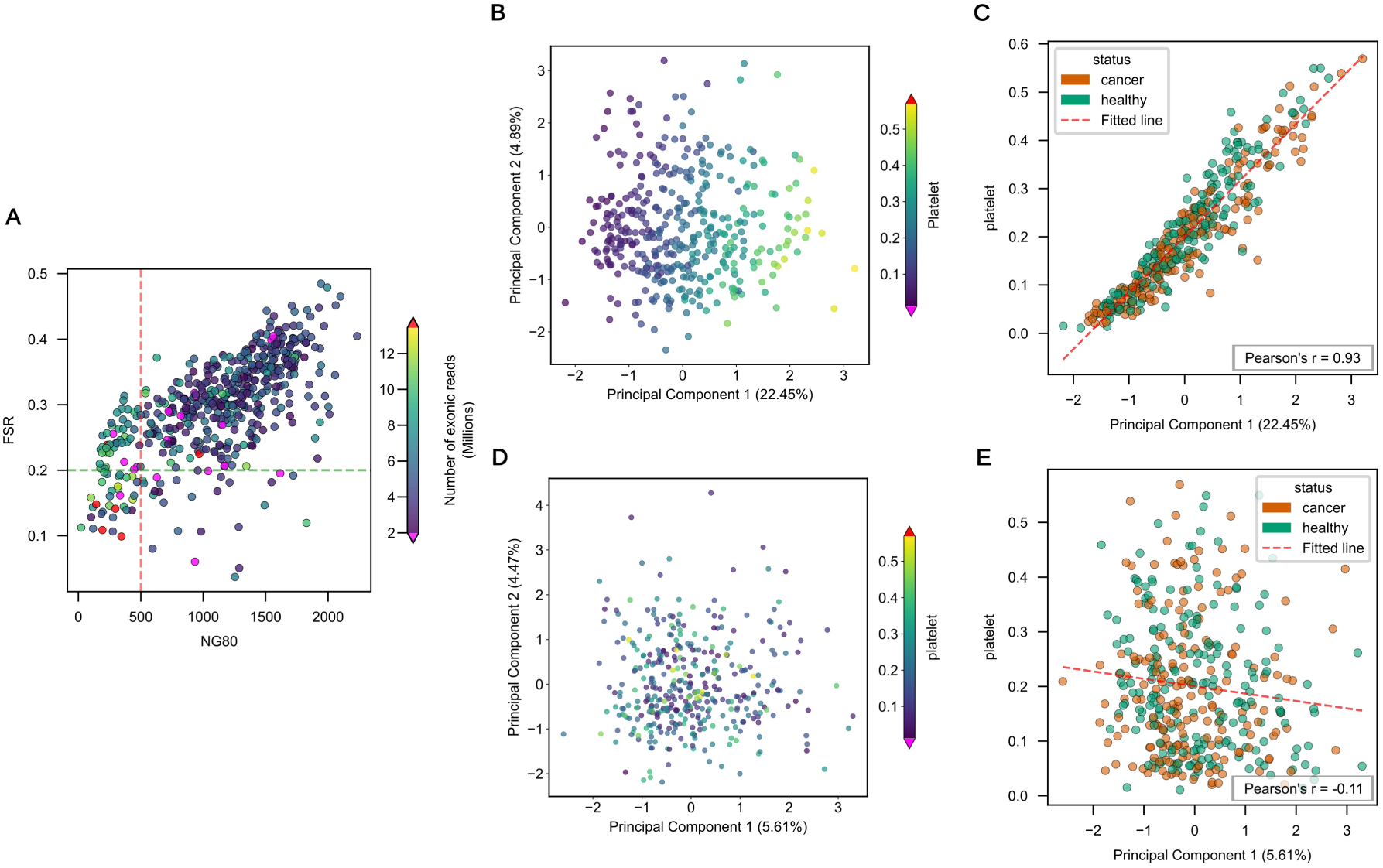
Quality-control assessment and correction of platelet-associated unwanted variation. **(A)** Quality-control filtering based on library complexity, splicing fraction, and exonic read depth. Each point represents an individual sample. NG80 indicates the number of genes accounting for 80% of mapped reads, and FSR indicates the fraction of spliced reads. Dashed lines show the QC thresholds for NG80 (>500) and FSR (>0.20). A third QC filter requires a minimum of 2 million exonic reads per sample and is represented by the color scale, with purple indicating samples below this threshold. Samples with very high exonic read counts, defined as more than 3 standard deviations above the mean, are shown in red for visualization purposes but were not excluded. QC filtering retained 388 of 505 samples. **(B-C)** Principal component analysis of the TMM-normalized count matrix and its association with platelet fraction. **(B)** Samples projected onto the first two principal components, with PC1 and PC2 explaining 22.45% and 4.89% of the variance, respectively. Points are colored by platelet fraction; purple and red indicate values more than 3 standard deviations below or above the mean, respectively. **(C)** PC1 from the TMM-normalized count matrix plotted against platelet fraction. Points are colored by sample status, and the red line indicates the fitted linear regression model for all samples. **(D-E)** Principal component analysis after RUVg correction using Platelet PanglaoDB genes as negative controls. **(D)** Samples projected onto the first two principal components of the RUVg-corrected count matrix, with PC1 and PC2 explaining 5.61% and 4.47% of the variance, respectively. Points are colored by platelet fraction as in panel B. **(E)** PC1 from the RUVg-corrected count matrix plotted against platelet fraction. Points are colored by sample status, and the red line indicates the fitted linear regression model for all samples.

### Technical and biological confounding effects in cfRNA-seq data

To visually assess the primary sources of variation in the high-quality dataset, we performed principal component analysis (PCA) on the TMM-normalized counts. We first verified that sample processing batches and sequencing runs were not associated with any of the first three principal components (Figure S2), suggesting the absence of laboratory-related technical batch effects. Consistent with previous findings (Moore *et al*., 2025; Nesselbush *et al*., 2025), we observed that the platelet fraction, derived via cell-type deconvolution, acts as a major driver of unwanted variability (Figure S3). In our TMM-normalized data, PC1 and PC2 accounted for 22.45% and 4.89% of the variance, respectively. Samples separated along the PC1 axis according to platelet fraction (Figure 2B). Indeed, platelet fraction was strongly correlated with the first principal component, showing a Pearson correlation of r = 0.93 (P ≤ 2.4 x 10^-175^) (Figure 2C). This strong positive relationship was consistent across both cancer and healthy control samples (cancer samples r = 0.94, P ≤ 2.1 x 10^-87^, healthy control samples r = 0.93, P ≤ 5.8 x 10^-93^), confirming that platelet contamination introduces a significant source of unwanted variation in cfRNA analyses.

To remove this platelet-driven noise, we followed the approach described by Nesselbush et al. (Nesselbush *et al*., 2025) and applied the RUVg normalization method using PanglaoDB platelet marker genes (Franzén, Gan and Björkegren, 2019) as negative controls. This approach estimates latent factors representing unwanted variation associated with platelet signal, enabling us to mathematically isolate and subtract these confounding effects from the data. PCA of the resulting RUVg-corrected count matrix demonstrated that the vast majority of unwanted variation associated with platelets had been removed (Figure 2D). There was no longer a clear association between the platelet fraction and PC1 or PC2, and the overall variance explained by PC1 and PC2 was substantially reduced to 5.61% and 4.47%, respectively. Furthermore, the previously strong correlation between PC1 and platelet fraction was eliminated, reducing the Pearson correlation to r = -0.11 (P ≤ 3.3 x 10^-2^) across the cancer and healthy control samples (cancer samples r = -0.02, P ≤ 0.8, healthy control samples r = -0.19, P ≤ 6.2 x 10^-3^) (Figure 2E). While this correction did not fully recover a distinct separation of the samples by phenotype (Figure S4), it successfully removed the primary confounding source of variation which improves our overall ability to identify genuine disease biomarkers in downstream analyses.

### Identification and biological significance of cfRNA biomarkers distinguishing CRC from healthy control samples

To identify robust biomarkers capable of distinguishing CRC patients from healthy individuals and supporting the development of a machine learning (ML)-based screening test, we used differential expression analysis as a feature-selection step. To prevent data leakage, this analysis was performed exclusively within the training cohort. The model accounted for platelet-associated technical variation using RUV factors and additionally adjusted for sex. Genes were ranked by false discovery rate (FDR), and the top 2,000 genes were retained as candidate features for the model development.

Overall, 1,370 genes are significantly associated with CRC status at an adjusted *p* value < 0.01 (Figure 3A). Among these, 35 are strongly upregulated in CRC, with log2 fold changes greater than 1, whereas one is strongly downregulated, with a log2 fold change below −1.

**Figure 3.**
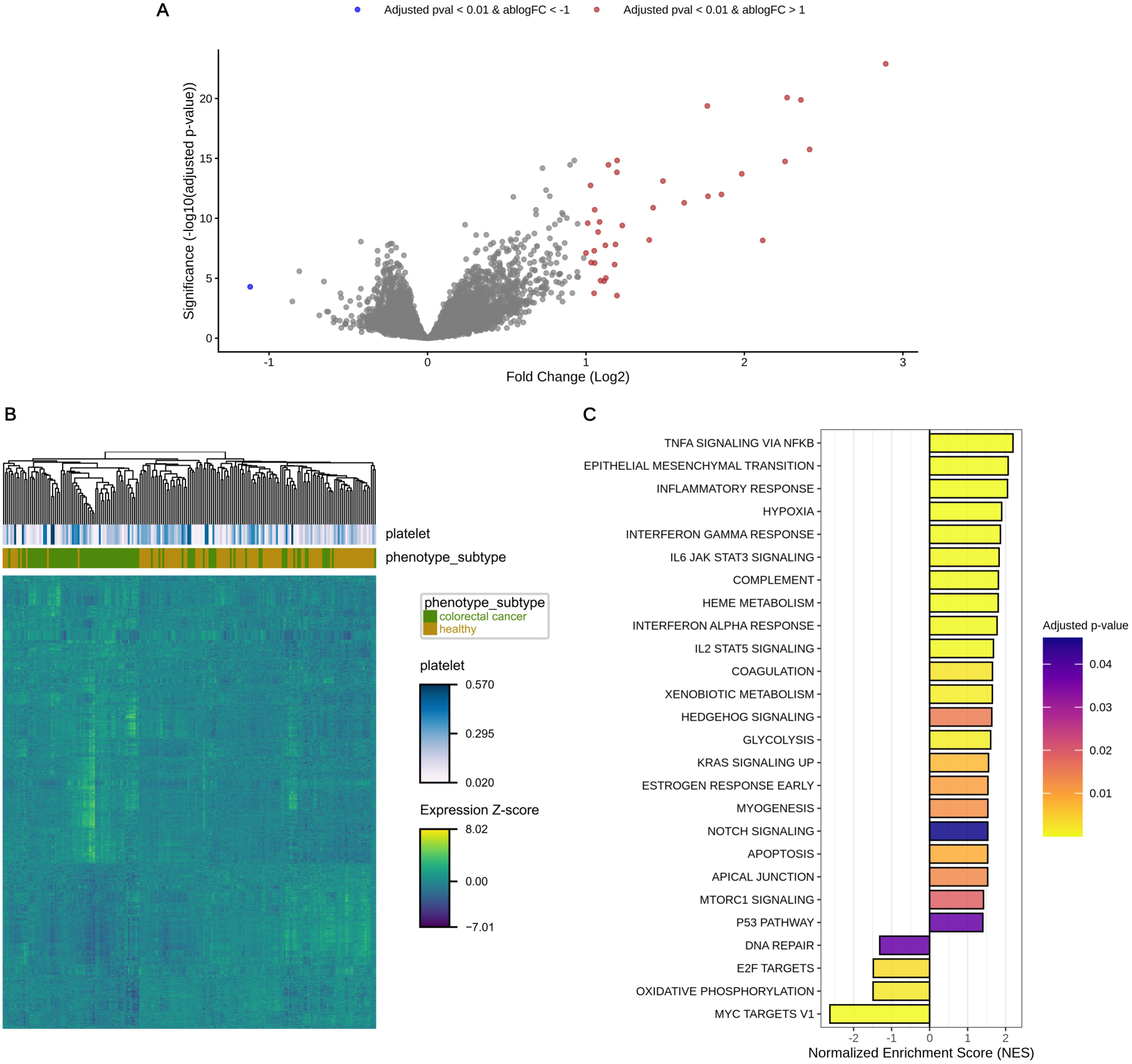
Differential expression–based feature selection and biological interpretation of plasma cfRNA in CRC. **(A)** Volcano plot of differentially expressed genes between CRC patients and healthy controls, showing log2 fold change versus −log10(adjusted p-value). Genes meeting the predefined thresholds of adjusted p-value < 0.01 and |log2 fold change| > 1 are highlighted as upregulated (red) or downregulated (blue) in CRC with respect to healthy controls, while non-significant genes are shown in grey. **(B)** Hierarchical clustermap of the top 2,000 differentially expressed genes ranked by adjusted P value. Rows represent genes and columns represent samples, with row-scaled Z-scores indicating relative expression levels. The top annotation bars represent sample metadata: *phenotype_subtype* defines the patient group (colorectal cancer in green and healthy controls in gold), while *platelet* indicates the predicted platelet contribution across samples, ranging from low (white) to high (blue). **(C)** Gene set enrichment analysis of FDR-ranked differentially expressed genes. Bars show normalized enrichment scores (NES) for the Hallmark gene sets with adjusted p-value < 0.05, with color indicating the significance level.

To evaluate expression patterns across these candidate features, we generated a hierarchical clustermap based on the list of the top 2,000 genes (Figure 3B). The expression profiles of these genes demonstrate structured patterns across samples, suggesting they capture meaningful biological variation rather than random noise. When clustering by samples, healthy control and CRC groups show substantial separation. Furthermore, annotating the clustermap with the original sample-associated platelet fraction reveals no evidence of platelet-driven clustering, indicating that platelet-related variation was effectively controlled and does not account for the observed sample structure.

Finally, to complement these gene-level results and evaluate whether the coordinated changes in gene expression are associated with known biological pathways, we performed Gene Set Enrichment Analysis (GSEA) (Figure 3C). This analysis identified a total of 26 significantly enriched Hallmark gene sets with an adjusted p-value lower than 0.05. 4 Hallmark gene sets are enriched in the genes downregulated in CRC, including gene sets implicated in oxidative phosphorylation and DNA repair. 22 Hallmark gene sets are enriched in the genes upregulated in CRC, including gene sets with roles in inflammation, hypoxia, epithelial–mesenchymal transition (EMT), coagulation, and metabolic function. Many signaling pathway-associated gene sets are also enriched in the genes upregulated in CRC, including the TNFA, IL-6/JAK2/STAT3, and Hedgehog signaling pathways.

### Colorectal cancer model reaches 67% sensitivity at 90% specificity on the validation cohort

Using the platelet-corrected cfRNA-seq gene expression data we developed a machine learning model to classify samples between “healthy control” and “CRC”. To minimize the risk of overfitting and ensure model robustness, the model was trained only on the previously defined list of the top 2,000 DEGs. We then used 4-fold cross-validation (CV) within the training cohort for hyperparameter tuning, thereby preventing data leakage (Figure tmp1D). The trained classifier effectively discriminated between healthy control and CRC samples, achieving a mean AUC of 0.92 on the 4 CV evaluation folds, a mean sensitivity of 77% and a mean specificity of 94% (Figure 4A). The sensitivity was stable across cancer stages (Figure 4C), with mean across folds ranging from 75% (stage I) to 82% (stage IV). The number of samples in each evaluation fold during CV was larger for stage III (n = 15 samples per evaluation fold in average) compared to the other stages (n = 2 for stage I, n = 8 for stage II, n = 7 for stage IV), reflecting the sample distribution across cancer stages in the training cohort. The sample size was particularly small for stage I cancer samples, with only 6 samples in fold #1, 2 samples in fold #2, 1 sample in fold #3, and 1 sample in fold #4. Thus, the larger variability of the sensitivity for stage I cancer samples can be explained by the smaller number of samples available. The confusion matrix (Figure 4B) revealed that 91.6% of healthy control samples and 77.1% of cancer samples were correctly identified in the well-balanced training cohort (143 healthy control samples and 131 cancer samples). Among the incorrectly classified samples, more cancer samples were predicted as healthy control (false negatives) than healthy control samples were predicted as cancer (false positives), reflecting the model tuning for a high target specificity of 90%.

**Figure 4.**
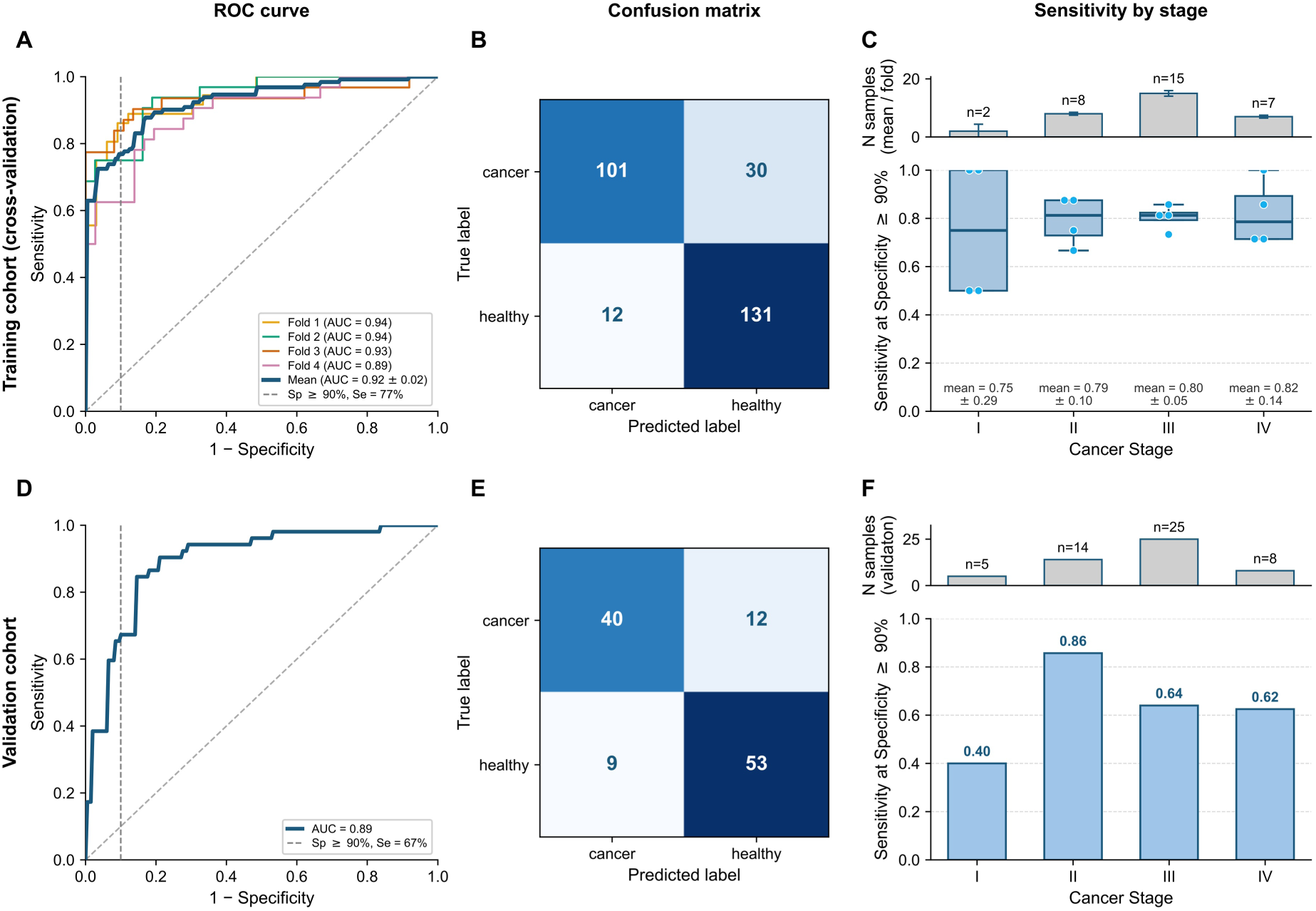
Cancer-versus-healthy classification performance in the training and validation cohorts. The top row (A–C) shows training performance of the ML model estimated by 4-fold cross-validation; the bottom row (D–F) shows performance on the held-out validation cohort. **(A)** Receiver-operating-characteristic (ROC) curves for the training cohort. Thin coloured lines are the four cross-validation folds (with their individual AUCs); the dark blue line is the mean ROC across folds (mean AUC = 0.93 ± 0.02). The dashed vertical line marks the 90% specificity operating point, at which the mean sensitivity is 77%. **(B)** Confusion matrix for the training cohort, pooled across the four held-out cross-validation folds (n = 274 samples: 131 cancer, 143 healthy). **(C)** Sensitivity at ≥ 90% specificity, stratified by cancer stage (I–IV), for the training cohort. Top: number of cancer samples per stage (mean per fold, error bars show SD). Bottom: per-fold sensitivity (box = interquartile range and median, points = individual folds); the per-stage mean ± SD is annotated below each box. **(D)** ROC curve for the held-out validation cohort (AUC = 0.89); sensitivity at the 90% specificity operating point (dashed line) is 67%. **(E)** Confusion matrix for the held-out validation cohort (n = 114 samples: 52 cancer, 62 healthy). **(F)** Sensitivity at ≥ 90% specificity, stratified by cancer stage (I-IV) for the validation cohort. Top: number of cancer samples per stage. Bottom: sensitivity per stage. AUC, area under the ROC curve; Se, sensitivity; Sp, specificity; SD, standard deviation.

We then evaluated the performance of our model on the held-out validation cohort. The model achieved an AUC of 0.89, with a sensitivity of 67% at a specificity threshold of 90% (Figure 4D-E). When analyzing performance across cancer stages (Figure 4F), the model demonstrated varying sensitivities within early-stage colorectal cancer, showing 40% for Stage I (n=5) and a notably higher 86% for Stage II (n=14), achieving a pooled early-stage sensitivity of 73.7% (Figure S5). For late-stage disease, the model achieved a combined sensitivity of 63.6% across Stages III (n=25) and IV (n=8).

## Discussion

In this study we compared the gene expression of healthy individuals and CRC at the cfRNA level to identify DEGs and develop an ML classifier for the binary classification of “healthy control” and “CRC”. To investigate if the DEGs that we identified are associated with biological processes implicated in CRC, and to confirm that our ML classifier is based upon real biological signals and mirrors known biology, we performed a GSEA and identified 26 enriched gene sets, 22 upregulated and 4 downregulated in CRC, with many of them having a known association with CRC as discussed below.

Inflammation is a key driver of CRC (Schmitt and Greten, 2021; Zhang and Qiao, 2022) and many of the most significantly enriched gene sets that we identified are associated with inflammation and CRC: TNFA signalling via NFKB (Mao, Zhao and Sun, 2025), inflammatory response (Ullman and Itzkowitz, 2011), hypoxia (Biddlestone, Bandarra and Rocha, 2015), interferon alpha and gamma response (Kamal *et al*., 2019; Liu *et al*., 2022), and IL-6-JAK-STAT3 signaling (Huang, Lang and Li, 2022). In addition, TNFα-NF-κB and IL-6-JAK-STAT3 signaling are also well-documented drivers of CRC pathogenesis that promote proliferation and anti-apoptotic programs (Lin *et al*., 2020; Berkovich *et al*., 2022). Detecting multiple high-confidence, inflammation-associated gene sets demonstrates that we are capturing inflammation-derived cfRNA signals in the blood of CRC patients.

In CRC epithelial to mesenchymal transition (EMT) has a central role in metastasis and progression (Lu, Kornmann and Traub, 2023; Nie *et al*., 2025). The EMT gene set is the second most significantly enriched gene set in our analysis, indicating that in CRC there is a strong EMT-derived cfRNA signal in the blood.

Interestingly we observe that glycolysis-associated genes are upregulated while oxidative phosphorylation-associated genes are downregulated in CRC. This suggests that the Warburg effect is being detected at the cfRNA level as the tumour cells shift from mitochondrial respiration toward aerobic glycolysis, as is known to occur in CRC (Pang and Wu, 2025).

We also identified a DNA repair-associated gene set that is significantly downregulated in CRC. Defective DNA repair is common in CRC, particularly in the mismatch repair and homologous recombination pathways (Wheeler *et al*., 2000; Moretto *et al*., 2022).

The enrichment of numerous gene sets central to CRC pathogenesis provides strong evidence that our cfRNA-based approach captures genuine biological signals rather than technical noise. The convergence of our findings with established CRC biology also validates the functional relevance of the DEGs employed for downstream feature selection and machine learning classification.

Using the gene expression data from the training cohort we built a machine learning model which achieved an AUC of 0.89 in the validation cohort. This performance is similar to the prediction performance obtained in other cfRNA studies. For instance, Chen et al. demonstrated the diagnostic value of combined human and microbe-derived plasma cfRNAs, achieving an AUC of approximately 0.90 for distinguishing cancer patients from healthy donors (Chen *et al*., 2022). In the validation cohort our model achieved 40% sensitivity for Stage I colorectal cancer and 86% for Stage II. While Stage I sensitivity is lower than the 72.1% reported in a similar study (Momen-Roknabadi *et al*., 2025), this discrepancy is likely driven by our limited number of Stage I samples (n=5). Notably, the performance of our model in Stage II demonstrates significant diagnostic potential. When compared to the ECLIPSE trial, where the sensitivity of the FDA-approved Shield™ cfDNA assay falls to 65% for Stage I lesions (Chung *et al*., 2024b), our results highlight the competitive utility of our cfRNA platform. These findings suggest that as stage-balanced cohorts expand, cfRNA will provide a robust orthogonal layer for early-stage detection, overcoming the sensitivity limitations inherent to DNA-based methods.

As mentioned above, the cfRNA transcriptome is profoundly sensitive to pre-analytical variables, such as sample handling, blood collection tube type, and centrifugation protocols (The exRNAQC Consortium *et al*., 2025; Tuñí-Domínguez *et al*., 2026). These technical factors frequently dominate the transcriptomic signal, introducing inter-laboratory batch effects that can mask genuine biological variation and complicate cross-study comparisons. Our study underscores the necessity of stringent standardization to overcome these hurdles. By implementing a uniform, optimized experimental protocol—including double-spin plasma isolation and double DNase treatment—we minimized the introduction of technical noise, allowing us to generate high-quality cfRNA profiles that are less susceptible to the batch effects identified in earlier literature.

Beyond experimental standardization, our results emphasize the importance of rigorous quality control (QC) as an essential safeguard for downstream biomarker discovery. In our dataset, approximately 23% of samples failed our QC criteria, which was based on NG80 and FSR. While the exclusion of these samples reduces the effective cohort size, it is a critical step to ensure that the classifier is trained on reliable biological signals rather than technical artifacts. We propose that these specific metrics serve to standardize sample eligibility and enhance the reproducibility of cfRNA investigations.

While our workflow scaled sample processing tenfold—from 50 in (Tuñí-Domínguez *et al*., 2026) to over 500 samples in this study—while maintaining high QC standards (proportion of samples passing QC filter 84% in Tuñi-Domínguez *et al*., 77% in this study), this retrospective study faces limitations that must be addressed to advance the platform toward clinical utility.

A key limitation is our sensitivity for early-stage CRC detection. Although we targeted a balanced distribution (65% early-stage versus 35% late-stage) retrospective recruitment resulted in an overrepresentation of late-stage samples, limiting our ability to capture subtle early-stage cfRNA signals. Future prospective studies must prioritize larger, stage-balanced cohorts, as our current Stage I sensitivity remains below the threshold required for clinical screening.

Furthermore, the retrospective nature of our sample collection imposes limitations on the granularity of clinical and pre-analytical metadata. In our retrospective framework, we lacked the detailed documentation of pre-analytical variables such as fasting status and patient lifestyle factors, which likely introduce residual noise that limits model precision. Future prospective studies will be essential to systematically capture these variables, allowing for more precise modeling and correction of unwanted variability.

In this work, we performed hierarchical clustering of the samples using gene expression data and observed that while samples mostly clustered according to phenotype, they were not perfectly separated (Figure 3B). This highlights the complex biological heterogeneity present in plasma cfRNA and underscores the necessity of employing an ML classifier that leverages multigene patterns rather than simple single-gene markers. Furthermore, while our application of RUVg normalization successfully mitigated platelet-driven noise, the residual heterogeneity observed in the PCA suggests that more sophisticated normalization strategies are required to achieve the precision needed for early cancer detection.

In summary, this study demonstrates that an optimized cfRNA profiling workflow, coupled with robust noise-correction strategies, can reliably overcome the technical challenges inherent to the cell-free transcriptome. By providing a dynamic, orthogonal readout of CRC tumour biology, our cfRNA platform effectively complements existing DNA-based methods. We envision that integrating cfRNA into multiomic strategies will offer a synergistic approach to improve detection sensitivity across all colorectal cancer stages, particularly in early-onset disease where the levels of ctDNA are very low. Looking ahead, the predictive accuracy of cfRNA-based classifiers will be further refined by the inclusion of larger, prospective, stage-balanced cohorts, the implementation of standardized preanalytical protocols, and the integration of orthogonal feature sets directly derived from cfRNA-seq data, such as somatic variants, circulating microbial signatures, gene fusions, and circular RNAs. As these cfRNA profiles evolve through more comprehensive data and multi-layered feature integration, they establish a clear path toward scalable frameworks for multi-cancer early detection.

## Methods

### Human participants and cohorts

All samples analysed in this study were collected with informed consent using protocols approved by the ethical and scientific committees at their respective centres. Retrospective samples and associated clinical data were obtained from Biobanco del Sistema Sanitario Público de Andalucía (Granada, Spain) and the Hospital Clinic Barcelona - Institut d’Investigacions Biomèdiques August Pi i Sunyer Biobank (Barcelona, Spain). All samples were processed following standard operating procedures with the appropriate approval of the Ethics and Scientific Committees. In accordance with the international ethical and legal framework, the release of biological samples and associated data from the HCB-IDIBAPS biobank is approved by both the internal and external Scientific Committees, as well as by the Ethics Committee from HCB. The IDIBAPS release was granted approval code A3-C22016. BBSSPA sample release was approved by the Portal de Ética de la Investigación Biomédica de Andalucía (PEIBA) with codes 2042-N-22 and SICEIA-2024-002798.

All participants were over the age of 40. If potential participants had been previously diagnosed with cancer then they must have been cancer-free for at least 10 years for inclusion in this study. Samples were collected from participants with CRC after diagnosis but before starting any type of treatment.

### Plasma isolation

Blood samples were collected through standard venipuncture in EDTA tubes which were then inverted 8 to 10 times for homogenization. The blood was processed to plasma within 4 hours of collection using a double-spin centrifugation protocol to minimise platelet content. The tubes were centrifuged at 1500x *g* for 10 minutes. The upper phase was carefully transferred to a new tube without disturbing the interphase and centrifuged at 2500x *g* for 15 min. Aliquots of the platelet-poor plasma were made and stored at -80°C.

### Plasma sample processing and sequencing

1 ml of each plasma sample was thawed on ice and centrifuged at 400x *g* for 2 minutes at 4°C. The supernatant was transferred to a new tube and 2 μl was taken for haemolysis quality control using a Nanodrop One (Thermofisher) by measuring absorbance at 414 nM.

External RNA Controls Consortium (ERCC) RNA Spike-In Mix (Thermofisher) was diluted with nuclease-free water, fragmented at 94°C for 2 minutes, and then immediately placed on ice. 1.5 pg of fragmented diluted ERCC spike-ins was added to each 1 ml of plasma after the lysis step during cfRNA isolation.

cfRNA was isolated from plasma using the Maxwell miRNA Plasma and Serum Isolation kit and Maxwell RSC 48 instrument (both Promega) according to the manufacturer’s instructions including the bovine DNase I treatment step. The eluate was concentrated using the Oligo Clean & Concentrator kit (Zymo Research) according to the manufacturer’s instructions with the RNA being eluted from the column using 9 μl of nuclease-free water. 8 μl of concentrated RNA was digested with HL-dsDNase (ArcticZymes Technologies) and then used as input for library preparation using the SMARTer Stranded Total RNA-Seq Kit v3 - Pico Input Mammalian (Takara Bio) according to the manufacturer’s protocol. Library concentration was measured using the Qubit 1X dsDNA High Sensitivity kit and Qubit Flex Fluorometer (both Thermofisher). Fragment size distribution was assessed on a TapeStation 4200 with the TapeStation HS D1000 kit (both Agilent Technologies). For sequencing, libraries were quantified by qPCR using the NGS library quantification kit (Takara Bio). Equimolar-pooled libraries were sequenced on the Illumina NextSeq2000 platform to generate 150 base pair paired-end reads.

Plasma samples underwent RNA extraction and library preparation in batches of 16 or more and were subsequently sequenced in batches of 60. To mitigate potential technical batch effects, samples were systematically distributed across all processing and sequencing batches to ensure a balanced representation of phenotypes and cancer stages.

### Primary and secondary bioinformatics analysis

All RNA-Seq datasets were processed using the nf-core/rnaseq (Patel *et al*., 2022) Nextflow (Di Tommaso *et al*., 2017) pipeline (version 3.8.1) from raw data (FASTQ files), using STAR (Dobin *et al*., 2013) as the read mapping software and Salmon (Patro *et al*., 2017) to quantify annotated transcripts. All samples were processed using the same custom STAR parameters (“--readFilesCommand zcat --twopassMode None --outSAMprimaryFlag OneBestScore --outFilterMismatchNmax 999 --outFilterMismatchNoverReadLmax 0.04 -- alignIntronMin 20 --alignIntronMax 1000000 --alignMatesGapMax 1000000 –outFilterType BySJout --alignSJDBoverhangMin 3 --alignSJoverhangMin 8 --peOverlapNbasesMin 40 --peOverlapMMp 0.8 --quantMode TranscriptomeSAM --quantTranscriptomeBan Singleend --outSAMattributes All”). Pipeline parameters were adapted to the cDNA library preparation method used in the dataset. Nextflow configuration files are available in the project’s GitHub repository (https://github.com/Flomics/fl-cfRNAmeta), under the nextflow/ subdirectory. We used the genome assembly version GRCh38 coupled with the GENCODE v39 annotation (Mudge *et al*., 2025). We incorporated the synthetic SIRV-set 3 spike-in transcripts (including the ERCC mix) to both the reference genome and annotation.

Mapped fragments were deduplicated using umiTools “UMI-dedup” (Smith, Heger and Sudbery, 2017). The fraction of spliced reads (FSR) was calculated as the fraction of uniquely mapped, deduplicated reads that span introns (i.e., with an “N” in their CIGAR string). The fraction of exonic reads (FER) was calculated as the fraction of STAR-mapped and deduplicated reads with at least one base of overlap with a GENCODE-annotated exon (excluding synthetic spike-in sequences).

### Data normalization

Isoform-level quantification performed by Salmon was summarized to gene-level counts using the tximport R package (Soneson, Love and Robinson, 2015). Lowly expressed genes were excluded by retaining genes with at least 10 reads in a sufficient number of samples, corresponding approximately to 70% of the smallest experimental group. The raw count matrix was normalized using the trimmed mean of M-values (TMM) method implemented in edgeR, to account for differences in library size and RNA composition across samples.

### RNA library diversity analysis

The NG80 metric (number of genes accounting for 80% of a sequencing library’s mapped and deduplicated reads) was calculated using the Salmon raw counts matrix as input, as produced by the standard nf-core/rnaseq pipeline. The full script for the calculation of NG80 can be found on the code repository (https://github.com/Flomics/fl-cfRNAmeta/blob/dev/src/ng80.R). Briefly, for each sample, genes were ranked by expression level (read counts) in descending order. The cumulative number of genes required to reach 1%, 5%, 10%, 50%, and 80% of total reads was calculated for each sample. This metric provides a good proxy into the complexity and heterogeneity of gene expression, with fewer genes needed indicating more concentrated expression in a subset of genes.

### RNA biotype classification

Based on Ensembl/GENCODE gene-biotype annotation (Mudge *et al*., 2025), a custom biotype classification was created to highlight relevant categories in the context of cfRNA. Labels for specific Ensembl biotypes were merged into more general categories: protein-coding transcripts were relabeled as “mRNA”, various pseudogenes subtypes combined under the category “Pseudogene”, related small RNAs such as snRNA and snoRNA were grouped together, whereas other small RNAs types (scaRNA, sRNA, scRNA and vault RNA) were classified into “Other small RNAs”. Finally, signal recognition particle RNAs (srpRNA, a subtype of misc_RNA consisting in genes RN7SL1, RN7SL2, and RN7SL3), mitochondrial ribosomal RNAs (MT rRNA), mitochondrial transfer RNAs (MT tRNA) and nuclear ribosomal RNA (rRNA) were categorized separately. The remaining categories were grouped under “Other RNA”. ERCC spike-ins were excluded from this analysis. A detailed mapping between the Ensembl biotypes and our custom biotype categories is provided in Supplementary table 1.

Gene-level TMM-normalized expression values were aggregated per biotype for each sample, by summing the TMM values of all genes belonging to each biotype category. For visualization and comparative analysis, aggregated gene expression values per biotype were converted to percentages, representing the proportion of total TMM values per sample attributed to each biotype. Mean values per biotypes across multiple samples in each experimental batch were calculated to facilitate comparisons.

### Deconvolution algorithm

The nu-SVR deconvolution method, as developed by Vorperian et al. (Vorperian, Moufarrej and Quake, 2022b), was employed to predict the relative cell type contribution in cfRNA. We used Tabula Sapiens 1.0 (Tabula Sapiens Consortium* *et al*., 2022), as provided in the Vorperian et al. paper, as the cell type basis matrix for nu-SVR.

### Platelet correction

To account for platelet-derived transcriptomic variation, we adopted a previously described correction strategy based on the removal of unwanted variation (RUVg) (Risso *et al*., 2014) using platelet genes as negative controls (Nesselbush *et al*., 2025). This approach models platelet-associated expression variability as an unwanted source of variation and estimates latent factors representing this signal while preserving biological variation of interest.

In brief, TMM-normalized expression values were log2-transformed using a pseudocount before correction. Platelet-associated variation was then estimated using the RUVg method implemented in the RUVSeq R package (Davide Risso [Aut, 2017), with platelet marker genes obtained from PanglaoDB used as negative controls. RUVg was applied to the log-transformed expression matrix, and the corrected normalized expression values returned by RUVSeq were retained for downstream analyses. The estimated factors of unwanted variation were also stored and used in the differential expression analysis.

This platelet-correction method was applied separately to the training dataset and to the validation dataset to avoid any information leakage.

### Dimensionality Reduction

Dimensionality reduction of gene expression data was performed using principal component analysis (PCA) implemented in the scikit-learn Python library (v1.7.2) (Pedregosa et al., 2011). To stabilize variance and reduce the influence of highly expressed genes, TMM-normalized expression values were log2-transformed with a pseudocount prior to PCA. PCA was then performed to identify the primary sources of transcriptomic variation across samples. The first two principal components, PC1 and PC2, were used to represent samples in a two-dimensional space and provide a global overview of the underlying structure of the dataset. Samples were colored according to relevant sample characteristics, technical variables, and quality-control metrics to visually assess their associations with the principal components In particular, the relationship between the PCA representation and the estimated platelet fraction obtained through deconvolution was evaluated to determine the extent to which platelet abundance contributed to the observed transcriptomic variation.

Similarly, PCA was performed on the platelet-corrected expression values, and the first two principal components were used to visualize the structure of the corrected dataset.

### Differential expression

Differential expression analysis was performed using the edgeR R package (Robinson, McCarthy and Smyth, 2010), based on TMM-normalized expression values. Gene-wise expression counts were modeled using negative binomial generalized linear models. To estimate expression differences associated with the individual’s phenotype, while controlling for potential confounding, the design matrix included sex and the platelet-associated factor of unwanted variation estimated using RUVg as covariates. The model was specified as: expression ∼ sex + platelet factor + phenotype. Gene-wise dispersions were estimated using the edgeR empirical Bayes framework, and differential expression was assessed by comparing healthy control vs colorectal cancer groups. P values were adjusted for multiple testing using the Benjamini–Hochberg false-discovery rate procedure (Benjamini and Hochberg, 1995). Genes with a false-discovery rate below < 0.01 and an absolute log2 fold-change greater than 1 were considered differentially expressed.

### Gene set enrichment analysis (GSEA)

Gene Set Enrichment Analysis (GSEA) was performed using a custom automated R pipeline developed in R (v4.1.2) (R Core Team, 2021) everaging the clusterProfiler suite of tools (v4.2.2) within the Bioconductor framework (Huber *et al*., 2015). Differential expression (DE) input genes were first filtered to map Ensembl identifiers to their respective NCBI Entrez Gene IDs using the org.Hs.eg.db Homo sapiens annotation package (v3.14.0) (Carlson, 2017); in instances where multiple transcripts mapped to a single Entrez ID, the entry with the highest absolute significance was retained. To capture both the magnitude and direction of expression changes, genes were ranked according to a metric calculated as −log10 (FDR)×sign(log2 Fold Change). For genes returning an FDR of exactly 0, the value was imputed using the minimum non-zero FDR present in the dataset to avoid infinite values during transformation. GSEA was executed across a comprehensive suite of functional ontologies. This evaluation encompassed Gene Ontology categories (Biological Process [BP], Cellular Component [CC], Molecular Function [MF] via clusterProfiler (Yu *et al*., 2012) and the GO.db metadata package v3.14.0), Reactome pathways (via ReactomePA v1.38.0 (Yu and He, 2015)), Disease Ontology networks (via DOSE v3.20.1 (Yu *et al*., 2015)), and Molecular Signatures Database (MSigDB v2024.1) collections (Hallmark [H] and C1 to C8), a joint project of the University of California San Diego and the Broad Institute (Subramanian *et al*., 2005; Liberzon *et al*., 2011, 2015). Across all collections, gene set size restrictions were strictly enforced to include only functional groups containing between 10 and 500 genes. Permutation-based enrichment statistics were derived using 10,000 simple permutations (nPermSimple), and the mathematical boundary parameter eps was explicitly set to 0. Statistical significance was defined using an adjusted p-value (FDR) threshold of ≤0.05. Highly enriched terms, semantic networks, and distribution profiles were visualized using enrichplot (v1.14.2) (Guangchuang Yu, 2018) and ggplot2 (v4.0.2) (Wickham, 2016).

### Machine learning training and evaluation

Binary classification was performed using the Extreme Gradient Boosting (XGBoost) algorithm (Chen and Guestrin, 2016), selected for its efficacy in modeling complex, high-dimensional transcriptomic data through integrated L1 and L2 regularization. To ensure robust model development and mitigate overfitting, 4-fold stratified cross-validation was implemented using scikit-learn (v1.7.2) (Pedregosa *et al*., 2011), maintaining a minimum of 30 samples per fold. Hyperparameter optimization was conducted via grid search; candidate models were ranked primarily by mean F0.25 score, with the area under the receiver operating characteristic curve (AUC) and the training computation time serving as tie-breakers. Upon optimal hyperparameter selection, the final classifier was trained on the complete training cohort. The optimized parameters were: grow_policy=’depthwise’, learning_rate=0.1, max_depth=3, n_estimators=400, and subsample=0.9. The classification threshold was calibrated on the training set to achieve a specificity ≥90% and subsequently applied to the independent validation cohort.

## Funding Information

The project ‘LiquiDx’ (reference CPP2021-008897) is financed by the Spanish Ministry of Science, Innovation and Universities (MCIN/AEI/10.13039/501100011033) and by the European Union “NextGenerationEU/PRTR”. C.T. and G.A. are supported by the Industrial Doctorates Plan of the Department of Research and Universities of the Government of Catalonia (references 2022-DI-00108 and 2024-DI-00013, respectively). M.W., P.S. and P.M. are supported by the Torres Quevedo programme (references PTQ2022-012611, PTQ2022-012612 and PTQ2023-012932, respectively), which is financed by the Spanish Ministry of Science, Innovation and Universities (MCIN/AEI/10.13039/501100011033). We acknowledge support of the Spanish Ministry of Science and Innovation through the Centro de Excelencia Severo Ochoa (CEX2020-001049-S, MCIN/AEI/10.13039/501100011033), and the Generalitat de Catalunya through the CERCA programme.

## Disclosures

P.M., L.S., G.A., B.N., C.T.-D., L.C., P.S., M.W., L.G., E.C.R., S.P.-F., and J.L. are or were employees of Flomics Biotech SL.

## Data Availability

Summary data are included in the article and supplementary materials. Due to commercial confidentiality, raw sequencing and expression matrices are not publicly deposited. Processed data may be requested from the corresponding author for academic validation, subject to a formal data-sharing agreement.

## Acknowledgements

We are indebted to the HCB-IDIBAPS Biobank for sample and data procurement. We thank the partners of the LiquiDx project for scientific advice and sample and data procurement: Health and Biomedicine, Leitat Technological Center, Barcelona Science Park, Barcelona 08028, Spain and the Andalusian Public Health System Biobank, Coordinating Node, 18016 Granada, Spain. We are grateful to the CRG Core Technologies Programme for their support and assistance in this work. We are grateful to João Curado and Esther Lizano for their scientific input and reviewing of the manuscript.

## Supplementary materials

### Supplementary figures

Provided as a separate document, with legends.

### Supplementary tables

Supplementary table 1

Detailed mapping between the Ensembl biotypes and our custom biotype categories.

## Notes

### Author Declarations

Ethics committee of Hospital Clínic de Barcelona - August Pi i Sunyer Biomedical Research Institute Biobank (HCB-IDIBAPS Biobank) gave ethical approval for this work (A3-C22016). Ethics committee of Portal de Ética de la Investigación BiomÉdica de Andalucía (PEIBA) gave ethical approval for this work (2042-N-22 and SICEIA-2024-002798).

