## Supplementary Figures for "A Robust Cell-Free RNA Approach for the Early Detection of Colorectal Cancer"

#### Figure S1

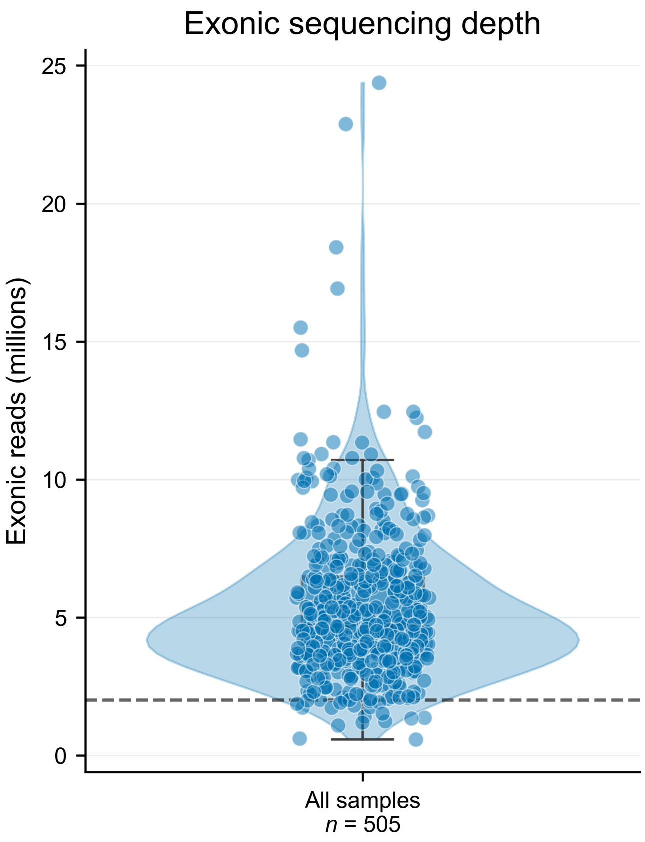


**Figure S1. Exonic sequencing depth across all samples.**

Violin plot showing the distribution of exonic read counts for all samples in the dataset (n=505 samples), expressed in millions. Points represent individual samples. The embedded boxplot shows the median and interquartile range (IQR), with whiskers extending to 1.5 × IQR. The dashed horizontal line indicates the predefined 2M minimum sequencing-depth QC filter.

### Figure S2

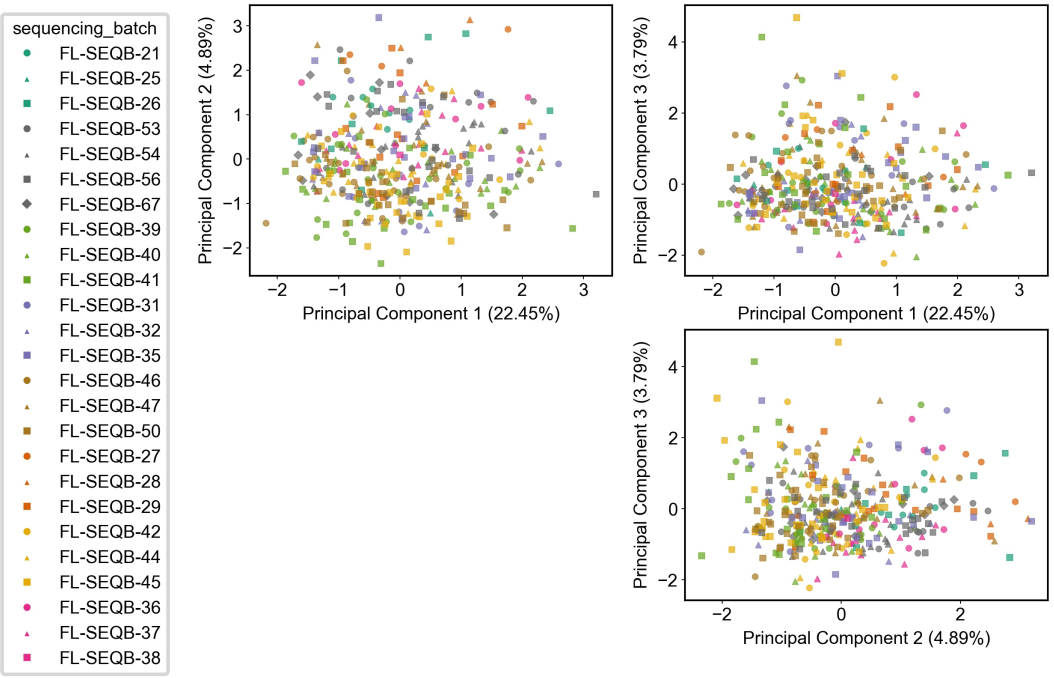


**Figure S2. High-quality standardized experimental processing minimizes batch-associated variation in cfRNA profiles.**
Principal component analysis (PCA) of cfRNA profiles generated using the standardized experimental workflow, including double-spin plasma isolation and double DNase treatment. Each point represents one sample, with color and shape indicating the individual sequencing batch. Samples were intentionally distributed across processing and sequencing batches to minimize confounding between phenotype and laboratory batch effects. Samples from different batches are broadly interspersed across the first principal components, with no evident batch-specific clustering. This extensive mixing supports the consistency and technical quality of sample processing and indicates that sequencing batch is not a major driver of variation in the resulting cfRNA profiles.

### Figure S3

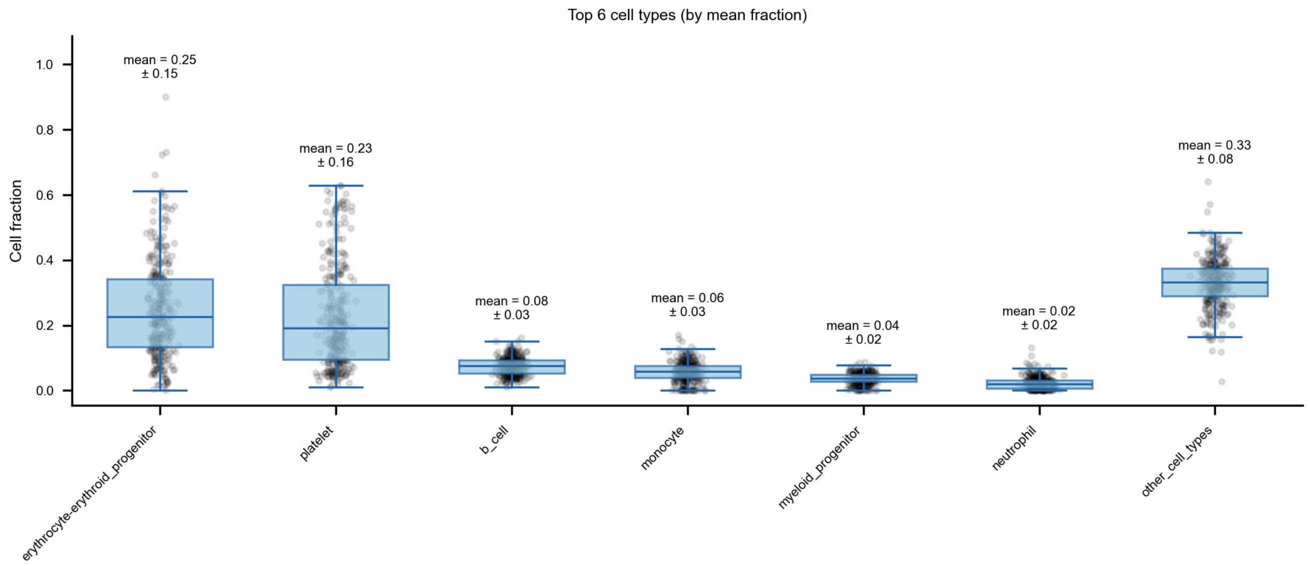


**Figure S3. Predicted cell types of origin contributing to the cell-free transcriptome.**

Relative abundance (cell fraction) of the top six predicted cell types of origin contributing to the cell-free transcriptome across the studied cohort. Individual data points represent independent plasma samples. Boxplots indicate the median (horizontal blue center line) and interquartile range (IQR), with whiskers extending to the maximum and minimum values excluding outliers. Above each boxplot, text annotations denote the computed mean fraction and standard deviation (±SD) for each respective group. Erythrocyte-erythroid progenitor and platelet constitute the predominant sources of circulating transcripts. All remaining cell types aggregated under "Other cell types".

### Figure S4

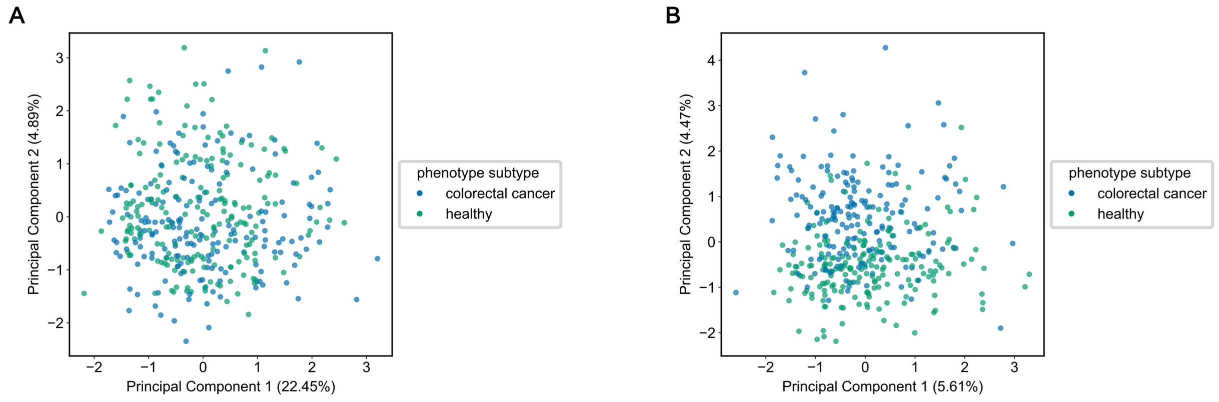


**Figure S4. Principal component analysis before and after correction of platelet-associated unwanted variation with RUVg showing distribution by phenotype.**
**(A)** Samples projected onto the first two principal components of the TMM-normalized count matrix, with PC1 and PC2 explaining 22.45% and 4.89% of the variance, respectively.
Points are colored by phenotype.

**(B)** Samples projected onto the first two principal components after RUVg correction using Platelet PanglaoDB genes as negative controls, with PC1 and PC2 explaining 5.61% and 4.47% of the variance, respectively.
Points are colored by phenotype.

### Figure S5

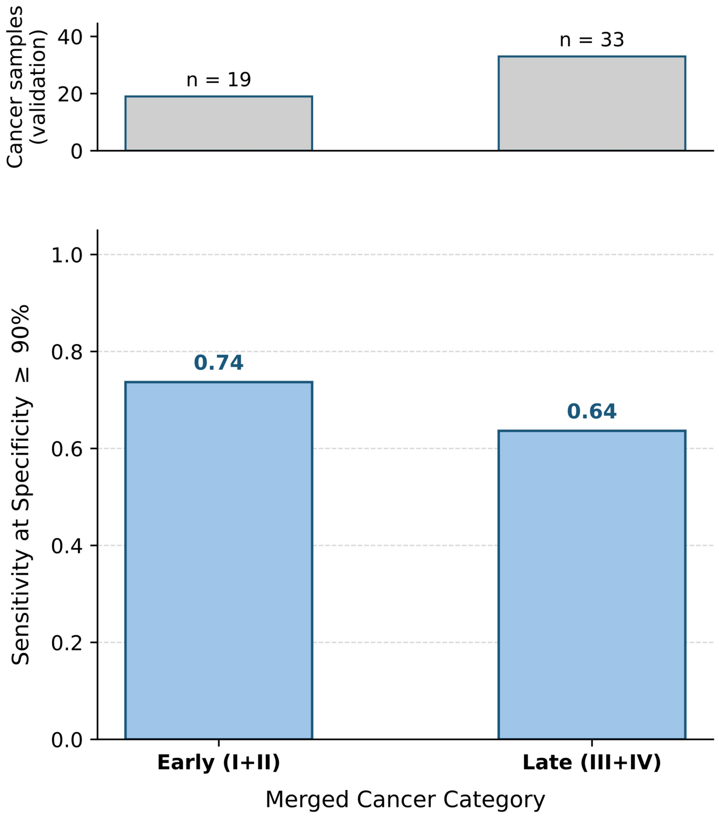


**Figure S5. Sensitivity of the classifier on the held-out validation set stratified by merged cancer stages.**

An XGBoost classifier was evaluated to distinguish colorectal cancer from healthy plasma samples using the top 2,000 differentially expressed features and platelet-corrected cfRNA-seq gene expression data. The figure shows the validation performance across grouped clinical classifications. Top: Absolute number of cancer samples per merged category in the held-out validation set (n=52 total cancer samples). Bottom: Classification sensitivity evaluated at the fixed ≥90% specificity operating point, stratified by merged cancer category into Early (Stages I and II combined; n=19) and Late (Stages III and IV combined; n=33). Exact sensitivity values are annotated in bold text above each bar.
